# Psychosocial factors affecting COVID-19 vaccine uptake in the UK: a prospective cohort study (CoVAccS – wave 3)

**DOI:** 10.1101/2022.03.25.22272954

**Authors:** Louise E. Smith, Julius Sim, Megan Cutts, Hannah Dasch, Richard Amlôt, Nick Sevdalis, G James Rubin, Susan M. Sherman

**Author notes:** Corresponding author: Louise E Smith, Post-doctoral Researcher. Department of Psychological Medicine, King’s College London, Weston Education Centre, Cutcombe Road, London, SE5 9RJ. Twitter handle: @louisesmith142.

## Abstract

**Background:** We investigated factors associated with COVID-19 vaccine uptake, future vaccination intentions, and changes in beliefs and attitudes over time.

**Methods:** Prospective cohort study. 1500 participants completed an online survey in January 2021 (T1, start of vaccine rollout in the UK), of whom 1148 (response rate 76·5%) completed another survey in October 2021 (T2, all UK adults offered two vaccine doses). Binary logistic regression analysis was used to investigate factors associated with subsequent vaccine uptake. Content analysis was used to investigate the main reasons behind future vaccine intentions (T2). Changes in beliefs and attitudes were investigated using analysis of variance.

**Findings:** At T2, 90·0% (95% CI 88·2%-91·7%) of participants had received two doses of a COVID-19 vaccine, 2·2% (95% CI 1·3%-3·0%) had received one dose, and 7·4% (95% CI 5·9%-8·9%) had not been vaccinated. Uptake was associated with higher intention to be vaccinated at T1, greater perceived vaccination social norms, necessity of vaccination, and perceived safety of the vaccine. People who had initiated vaccination reported being likely to complete it, while those who had not yet received a vaccine reported being unlikely to be vaccinated in the future. At T2, participants perceived greater susceptibility to, but lower severity of, COVID-19 (*p*<0.001), than T1. Perceived safety and adequacy of vaccine information were higher (*p*<0.001).

**Interpretation:** Targeting modifiable beliefs about the safety and effectiveness of vaccination may increase uptake.

**Funding:** Data collection was funded by a Keele University Faculty of Natural Sciences Research Development award and a King’s COVID Appeal Fund award.

**Research in context:** *Evidence before this study:* COVID-19 vaccination intention was high at the start of the vaccine rollout in the UK. Research suggests that psychosocial factors are associated with vaccine uptake. However, most research on uptake of the COVID-19 vaccine has investigated factors associated with vaccination intention, and used a cross-sectional design.

*Added value of this study:* We used a prospective cohort study (T1 conducted in January 2021, the start of the UK vaccine rollout; T2 conducted in October 2021, all UK adults offered two vaccine doses) to investigate factors associated with subsequent COVID-19 vaccination. Qualitative data on the main supporting reasons for future vaccination intentions in those partially or not vaccinated were analysed using content analysis. Changes in vaccine beliefs and attitudes (generally and COVID-19 specific) were also analysed.

*Implications of all the available evidence:* In our sample, more people reported having been vaccinated than had previously reported intending to be vaccinated. Vaccine uptake was strongly associated with previous vaccination intention, perceived social norms of vaccination, and greater perceived necessity and safety of vaccination. Those who had received at least one COVID-19 vaccine reported being likely to complete the schedule, whereas those who had not received a vaccine reported being unlikely to receive a vaccine.

## Introduction

One of the main lines of defence against COVID-19 has been vaccination. In the United Kingdom (UK), intention to receive a COVID-19 vaccine when one became available was reasonably high, with 74% indicating that they were likely to be vaccinated against COVID-19 in a previous survey conducted by our team in January 2021 (at the start of the vaccine rollout).^1^ Other UK studies have found comparable rates of intention to be vaccinated (63% to 89%).^2-4^ Differing rates can be explained by different timepoints in the pandemic and different questions used. On 19 July 2021, all UK adults had been offered a first dose of a COVID-19 vaccine.^5^ At this point, people were eligible for their second vaccine eight weeks after they had received their first, meaning that all UK adults would have been offered a full course by 13 September 2021.

Most studies investigating COVID-19 vaccination uptake have explored factors associated with intention to receive a vaccine using cross-sectional survey methods, finding that vaccination intention is associated with psychological, contextual and sociodemographic factors. In the UK, vaccination intention has been associated with: greater perceived necessity of the vaccine, lower perceived safety concerns, believing that others like you will be vaccinated (i.e. more supportive perceived social norms), and perceiving a low risk of infection.^1,2,4^ Not intending to be vaccinated has been associated with not having received an influenza vaccine last year and lower adherence to other Government guidelines.^1,4^ Sociodemographic factors associated with not intending to be vaccinated have included: lower income, lower education, belonging to a minoritized ethnic group, younger age, being female, and living with a dependent child.^4,6^

While these studies informed communication campaigns at the start of the vaccine rollout, there are known differences between intended and enacted health behaviours.^7^ To the best of our knowledge, there are very few studies investigating psychological and contextual factors (i.e. not sociodemographic factors) associated with COVID-19 vaccine uptake in the UK general population. Among UK healthcare workers, not having had a COVID-19 vaccine was associated with previous confirmed SARS-CoV-2 infection, as well as younger age, being female, greater deprivation, and belonging to a minoritized ethnic group.^8^

Globally, few longitudinal studies exist investigating vaccine uptake. One conducted in China found that previous vaccination intention (before the start of the vaccination campaign) and believing that the vaccine was safe were associated with vaccine uptake, whereas vaccine shortages were associated with not being vaccinated.^9^ In a study of students (aged 17 to 28 years) in the Netherlands, vaccination intention (when COVID-19 vaccines were approved but not yet available for young adults) was associated with later uptake.^10^ Greater worry (measured before COVID-19 vaccines were approved) was associated with vaccination intention, with mediation analyses indicating that there was an indirect effect of greater perceived severity of COVID-19 (measured before COVID-19 vaccines were approved) on uptake, through worry and vaccination intention. In Israel, vaccination intention (measured in the week before a COVID-19 vaccine was made available to the general public) was strongly associated with later behaviour (measured after vaccinations were available for all individuals).^11^ COVID-19 illness and vaccine attitudes and beliefs, perceived social norms, past influenza vaccination explained 86% of the variance in vaccination intention, which itself mediated associations with behaviour.

Beliefs about, and attitudes towards, COVID-19 vaccination are likely to have changed over the course of the pandemic, as vaccines were rapidly developed, tested, approved, and rolled out to the population. During the rollout, the AstraZeneca vaccine was linked to unusual blood clots with low blood platelets (published April 2021).^12^ This was the focus of widespread media attention and linked to the suspension of delivery of the vaccine in younger age groups in some countries.^13^ Pfizer and Moderna vaccines have also been linked to other very rare adverse effects (myocarditis and pericarditis), although these received less media attention.^14^ Research conducted in the United States (US) between March and August 2020 indicated that vaccination intention and general vaccine attitudes became more negative.^15^ However, since the start of the rollout, studies indicate more positive vaccine intentions and sentiments, with vaccine refusal and delay decreasing between October 2020 and July 2021 in the US.^16^ In Italy, more people agreed that vaccines were important to public health and fewer endorsed the idea that vaccines were created to make money for pharmaceutical companies in May 2021 compared to May 2020.^17^ In a cohort of UK older adults (aged 65 years and over), concerns about commercial profiteering and mistrust of vaccination decreased, while collective responsibility and worries about unforeseen future effects had increased.^18^

The aims of this study were to investigate: factors associated with subsequent uptake of a COVID-19 vaccine; changes in beliefs and attitudes about COVID-19, COVID-19 vaccination, and general vaccination beliefs and attitudes between January and October 2021; likelihood of further vaccination (completing or starting vaccine schedule in those partially or not vaccinated, respectively; and likelihood of accepting a booster vaccine); and reasons favouring or disfavouring future vaccination.

## Methods

This study reports data from the second and third rounds of the UK-wide ‘COVID-19 Vaccination Acceptability Study’ (CoVAccS), designated here as T1 and T2. Questions directed to parents about child vaccination are reported elsewhere.

### Design

This was a prospective cohort study. Participants completed an online survey at the start of the rollout of the COVID-19 vaccine in the UK (T1, 13–15 January 2021; results published in Sherman et al^1^) and after the vaccine had been offered to all adults (T2, 4–15 October 2021).

### Participants

Participants were eligible for the study if they were living in the UK, were aged 18 years or older, and had not completed round 1 of the CoVAccS study (data collected July 2020).^19^ Only participants who had taken part in round 2 of our survey^1^ (n=1500; T1, January 2021) were invited to take part in the third round of data collection (T2, October 2021) and formed the study cohort.

### Measures

Full survey materials are available online.^20^ To allow direct longitudinal comparisons, with the exception of demographic questions such as age and gender, the same questions were asked at T1 and T2.^1^ Further questions were added, as detailed below.

#### Uptake of vaccine

Participants were asked if they had been vaccinated against coronavirus. Response options were “yes, I’ve had one dose”, “yes, I’ve had two doses”, “no”, “don’t know” and “prefer not to say” (asked at T1 and T2). At T2, participants who reported they had been vaccinated were asked which vaccine they had received (choice of Pfizer-BioNTech, AstraZeneca, Moderna, Janssen [Johnson & Johnson], a made-up brand “Cambriona”, or another vaccine not listed above), to ascertain whether they had completed the full vaccine schedule. They were also asked if they would have preferred a different vaccine from the one they had received, and if so, which vaccine they would have preferred, using the same list.

The following questions were only asked at T2. Participants who reported that they had only had one dose were asked how likely they would be to have a second dose on an 11-point scale from “extremely unlikely” (0) to “extremely likely” (10), and to give the main reason why they were likely or unlikely to have a second dose. These questions were only asked to those who indicated that they had received a vaccine that needed two doses to be “fully vaccinated” (Pfizer-BioNTech, AstraZeneca, Moderna). Participants who had not been vaccinated were asked how likely they would be to get vaccinated using the same 11-point scale, and to give the main reason why they were likely or unlikely to have a vaccine.

We asked participants if they had had a COVID-19 booster vaccination. Those who indicated they had not had a booster were asked how likely they would be to have one if it became available to them.

#### Psychological and contextual factors

These questions were asked at both T1 and T2 and were informed by existing psychological theory and evidence on psychosocial factors affecting vaccination uptake.^1,19^ Participants were asked about the perceived risk of COVID-19 to themselves personally, to people in the UK, and to people in their local area (five-point scale from “no risk at all” to “major risk”). We also asked participants if they thought they had had, or currently had, a confirmed COVID-19 infection, and whether they personally knew anyone who had had COVID-19.

We measured participants’ beliefs and attitudes about COVID-19. At T1, eight questions, asking about perceived worry about catching COVID-19, perceived susceptibility to and severity of COVID-19, and the impact and management of COVID-19 were used. Questions were answered on an 11-point scale (“strongly disagree” [0] to “strongly agree” [10]).

Perceptions of vaccination were asked at T1 and T2. We measured general vaccine beliefs and attitudes using two items, asking about vaccination in general being a good thing and fear of needles. Beliefs and attitudes about COVID-19 vaccination were asked about using 21 questions, including perceived effectiveness of vaccination, social norms of vaccination, ease of vaccination, novelty and safety of vaccination, and (at T2) whether COVID-19 vaccination should be made mandatory. Questions were phrased to take into account whether the participant had already been vaccinated. All questions were answered using the same 11-point scale (“strongly disagree” [0] to “strongly agree” [10]).

At T1, participants were asked how likely they were to have a COVID-19 vaccination on an 11-point scale from “extremely unlikely” (0) to “extremely likely” (10).

#### Personal and clinical characteristics

Participants’ age, gender, ethnicity, religion, education, working situation, household income, and chronic illness status (self and household member if applicable) were collected at T1. As participants could have been diagnosed with a medical condition or changed job roles between rounds of data collection, we asked participants whether they had a chronic illness and about their current working situation at T2. We also asked participants if they had a vaccine for seasonal flu during the winter of 2020/2021.

### Ethics

Keele University’s Research Ethics Committee granted ethical approval for this study (reference: PS-200129).

### Analysis

#### Uptake of a COVID-19 vaccine at T2

We tabulated the association between categories of vaccination intention at T1 and subsequent vaccination uptake. These categories were designated a priori on the 0–10 scale as follows: 0–2 “very unlikely”, 3–7 “uncertain”, 8–10 “very likely.”^1^

Due to small numbers of participants who were partially vaccinated, we created a binary outcome variable (unvaccinated vs partially/fully vaccinated). We conducted a logistic regression analysis to investigate factors associated with subsequent uptake of COVID-19 vaccination. Explanatory variables were measured at T1, while vaccine uptake (outcome) was measured at T2. For these analyses, we excluded participants who reported that they had already been vaccinated against COVID-19 at T1 (*n*=30 at T1, *n*=24 at T2). Explanatory variables were entered into the regression analysis in two blocks, selected *a priori*.^1^ In the first block we entered vaccination intention, measured at T1. In the second block we added variables that had been significant predictors of vaccine intention at T1:^1^ four principal components representing i) social norms relating to vaccination, ii) perceived necessity of vaccination, iii) perceived safety of the vaccine, and iv) adequacy of information about the vaccine; an item indicating a belief that only those at risk of serious illness should be vaccinated; an item indicating that vaccination was just a way of vaccine manufacturers making money; and receipt of the influenza vaccine last/this winter (completed and intended behaviour combined to give a single binary item). The use of blocks allowed us to gauge the predictive strength of vaccination intention both before and after controlling for other potential predictors of vaccination status. The predictive strength of each model was calculated as the Tjur coefficient of discrimination;^21^ this statistic can take values between 0 and 1, with higher values indicating greater predictive power. Additionally, the goodness of fit of each model was measured as the deviance and the improvement in goodness of fit in the second model was tested through a likelihood ratio test on the model deviances. As the odds ratios for the predictors in the analysis could not be compared for their magnitude, owing to the different scales on which these variables had been measured, we also calculated standardized coefficients for each predictor.^22^ For the regression analysis, statistical significance was set at *p* ≤·05.

#### Changes in beliefs and attitudes about COVID-19 illness and vaccination between January and October 2021

In our previous analyses of T1 data, we used principal components analysis to summarise items relating to beliefs and attitudes about COVID-19 (four resulting components) and COVID-19 vaccination (five resulting components; see Sherman et al. 2022^1^ for more details). We assessed changes in beliefs and attitudes about COVID-19, COVID-19 vaccination, and general vaccine beliefs and attitudes between T1 (January 2021) and T2 (October 2021) using repeated measures ANOVA. Effect sizes were calculated as Cohen’s *f*; a value of ·10 is considered to represent small effect, a value of ·25 a medium effect, and a value of ·40 a large effect.^23^ In view of the number of hypothesis tests performed, statistical significance was set at a more stringent *p* ≤·01 for these analyses.

#### Future vaccine intentions, and reasons behind intention

Participants’ intention to receive future COVID-19 vaccines – second dose in those partially vaccinated or any COVID-19 vaccine in those not vaccinated, and a booster vaccine (asked to all) – was categorized using *a priori* cut-points (0–2 very unlikely; 3–7 uncertain; 8–10 very likely).^1,19^

Open-ended answers about participants’ main reasons why they were likely or unlikely to accept future vaccination were analysed qualitatively through content analysis. An emergent coding approach was used, whereby codes were identified from the data.^24^ Content analysis was undertaken by two authors (MC and HD), starting with the coding framework generated from analysis of similar data that had been collected at T1 of the CoVAccS study.^1^ Statements were jointly coded by these authors; any difference in opinion was resolved through discussion to give a final set of codes. Codes were applied separately to intention to complete the initial vaccine schedule (receive a second dose in those partially vaccinated) and to initiate the COVID-19 vaccine schedule (in those not vaccinated). We report codes by intention to receive future vaccines (very unlikely, uncertain, very likely).

For analyses investigating intentions to receive a booster vaccine, we excluded those who reported already having had a booster (*n*=25, 2·2% of sample).

## Results

### Participant characteristics

Of the 1500 participants who had completed T1, 76·5% (*n*=1148) also completed T2. The mean (SD) age of respondents (recorded at T1) was 48·2 (15·1) years and 53·2% (*n*=611) were female. The majority (86·1%, *n*=988) were of white ethnicity. A higher percentage of participants who completed T2 were female than of those who did not complete T2 (53·2% versus 44·3%). Those completing T2 were also older (mean age 48·2 years versus 37·3 years), and more likely to be of white ethnicity (86·1% versus 80·1%). The mean vaccination intention score was also higher in those who completed T2 (8·3 versus 7·7). Participant characteristics for both timepoints are reported in detail in Supplementary Table 1.

### Uptake of a COVID-19 vaccine at T2

A large majority of participants (90·0%; 95% CI 88·1%, 91·6%, *n*=1033/1148) reported having received two doses of a COVID-19 vaccine, with a further 2·2% (95% CI 1·5%, 3·2%; *n*=25) reporting having had one dose; 7·4% (95% CI 6·0%, 9·1%; *n*=85) had not been vaccinated (0·3% [*n*=4] preferred not to say, 0·1% [*n*=1] did not know).

Most participants (56·6%, *n*=599/1058) reported having the AstraZeneca vaccine, followed by the Pfizer-BioNTech vaccine (39·4%, *n*=417). Few reported having the Moderna (3·4%, *n*=36), another vaccine not listed (0·3%, *n*=3), or the Janssen (0·1%, *n*=1) vaccines (0.2% did not know, *n*=2). No one selected the made-up brand “Cambriona”. A minority (12·4%, 95% CI 10·5%, 14·5%, *n*=131/1058) reported preferring a different vaccine to the one they had been given; 687 (64·9%; 95% CI 62·0%, 67·8%) did not prefer another vaccine; 240 (22·7%; 95% CI 20·3%, 25·3%) did not know. Of these, 90·1% (*n*=118/131) reported preferring to receive the Pfizer-BioNTech vaccine (5·3% Moderna, *n*=7; 2·3% Janssen, *n*=3; 1·5% AstraZeneca, *n*=2; 0·8% prefer not to say, *n*=1).

#### Factors associated with subsequently being fully vaccinated at T2

More participants had been vaccinated at T2 (October 2021) than had indicated being very likely to do so at T1 (January 2021; *n*=1030 vaccinated, compared to *n*=847 very likely; Table 1). Almost all participants (99·9%) who indicated that they were very likely to be vaccinated had been vaccinated. Of those who had previously stated they were very unlikely to be vaccinated, 39·8% had been vaccinated; 85·9% of those who were uncertain had been vaccinated.

**Table 1.** Association between vaccination intention at T1 (January 2021, using *a priori* cut points) and subsequent vaccination status at T2 (October 2021). Data are frequencies (%).

| Vaccination status,<br>October 2021 | Vaccination intention, January 2021 |  |  | Total |
| --- | --- | --- | --- | --- |
|  | Very unlikely | Uncertain | Very likely |  |
| Vaccinated | 37 (39.8) | 146 (85.9) | 847 (99.9) | 1030 |
| Unvaccinated | 56 (60.2) | 24 (14.1) | 1 (0.1) | 81 |
| Total | 93 (100) | 175 (100) | 848 (100) | 1111 |

Vaccination intention was strongly associated with vaccine uptake, with an odds ratio of 1·89 (95% CI 1·71, 2·09) and a coefficient of discrimination of ·443 (Table 2). Addition of the other predictors in the second block significantly improved the fit of the model (*χ*^2^=29·41, *df*=7, *p*<0·001) and raised the coefficient of discrimination to ·501. Vaccine intention remained a significant predictor, with a slightly lower odds ratio of 1·43 (95% CI 1·21, 1·68). Three of the components – social norms relating to vaccination, necessity of vaccination, and perceived safety of the vaccine – were also significant predictors, but of less strength, as indicated by the standardized beta coefficients.

**Table 2.** Results of the logistic regression model analysing associations with vaccination intention. The odds ratios indicate the increase or decrease in the odds of vaccination for a one-unit increase in the predictor variable. The model was based on 1111 cases with complete data.

| Predictor variable | Level | Odds ratio | 95% confidence interval | p value | Standardized beta | Coefficient of discrimination for model |
| --- | --- | --- | --- | --- | --- | --- |
| **Univariable model** |  |  |  |  |  |  |
| Vaccination intention | 0–10 | 1.887 | 1.708, 2.085 | <.001* | .570 | .443<sup>†</sup> |
| **Multivariable model** |  |  |  |  |  |  |
| Vaccination intention | 0–10 | 1.429 | 1.214, 1.681 | <.001* | .301 | .501<sup>#</sup> |
| Component 1: social norms | — | 1.639 | 1.164, 2.308 | .005* | .145 |  |
| Component 2: necessity of vaccination | — | 2.137 | 1.350, 3.383 | .001* | .229 |  |
| Component 3: perceived safety of the vaccine | — | 1.937 | 1.318, 2.846 | .001* | .192 |  |
| Component 4: adequacy of information about the vaccine | — | 1.233 | 0.908, 1.674 | .180 | .061 |  |
| Only people who are at risk of serious illness need to be vaccinated | 0–10 | 1.073 | 0.944, 1.220 | .282 | .063 |  |
| Widespread vaccination is just a way to make money for vaccine manufacturers | 0–10 | 0.937 | 0.825, 1.064 | .313 | -.050 |  |
| Had/will have a vaccination for influenza last/this winter | Yes | 1.580 | 0.693, 3.600 | .277 | .067 |  |

\* $p \leq .05$ ; <sup>†</sup> model deviance = 285.103; <sup>#</sup> model deviance = 255.689.

### Changes in beliefs and attitudes about COVID-19 illness and vaccination between January and October 2021

Compared to January 2021, in October 2021, participants perceived COVID-19 to be less severe and have a smaller impact on one’s life, but perceived their own vulnerability to COVID-19 as higher (Table 3). Participants had greater trust in COVID-19 management, perceived COVID-19 vaccination to be safer, and were more likely to perceive that they had adequate information about the vaccine, but were less likely to think that freedom from restrictions could be achieved through vaccination in October 2021. Participants were also less likely to agree that only those who are at risk of serious illness from COVID-19 need to be vaccinated and that the way that vaccines were given went against the manufacturers’ recommendation. Fear of needles also decreased between January and October 2021. Comparison of the mean change in attitudes between those who had been partially or fully vaccinated and those who had not been vaccinated indicates the extent of the difference in the rates of change between these sub-groups.

**Table 3.**
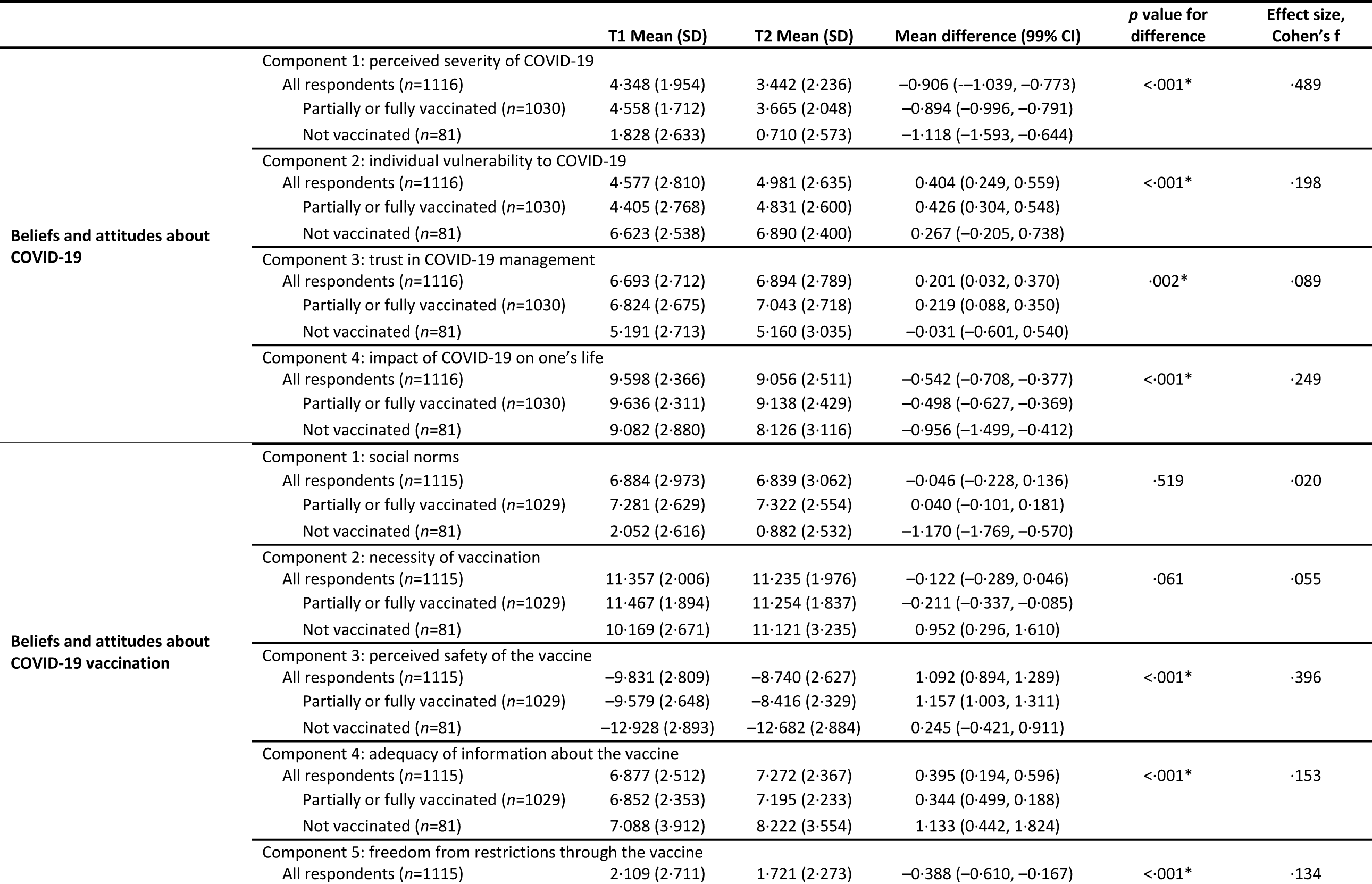
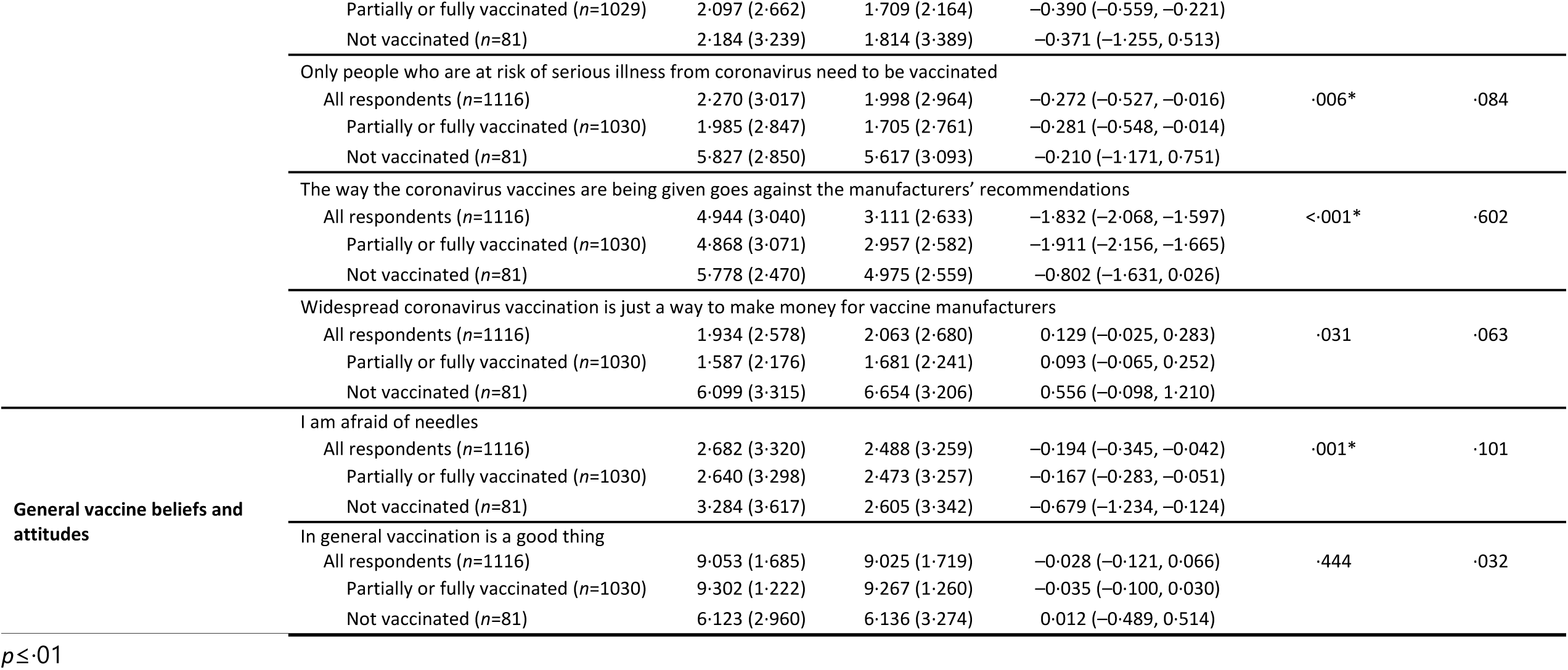
Changes in attitudes to COVID-19 between T1 and T2 (differences are T2 minus T1, i.e. October 2021 minus January scores; positive differences indicate a strengthening of attitude or belief from T1 to T2, negative differences a weakening from T1 to T2). Values are given for the sample as a whole and for those participants who subsequently did or did not vaccinate (5 participants did not report vaccination status). Those already vaccinated at T1 were excluded. To measure changes in the principal components between T1 and T2, the original component score coefficients from T1 were used to generate corresponding component scores at T2.

### Future vaccine intentions, and reasons behind intention

#### Receiving a second dose

Of 24 participants at T2 (October 2021) who had not completed their vaccine schedule (reported only receiving one dose), 79·2% (95% CI 59·5%, 90·8%, *n*=19) were very likely to have a second dose; 8·3% (95% CI 2·3%, 25·9%, *n*=2) were very unlikely, and 12·5% (95% CI 4·3%, 31·0%, *n*=3) were uncertain (see Supplementary Figure 1). The modal (most common) answer was the maximum value on the intention scale, with 63·5% participants (*n*=15) selecting “10 (extremely likely)”.

The most common reasons for having a second dose were to protect oneself, to be able to move about freely, and to protect others (Table 4). Lack of trust in authorities formed the main reasons for not being likely to have a second dose.

**Table 4.** Thematic categorization of codes generated by content analysis of reasons for or against having a second dose of the COVID-19 vaccine, by vaccination intention at T1. Data are the frequency with which codes were identified and themes are presented in descending order of overall frequency.

| Theme | Codes | Vaccination intention |  |  |
| --- | --- | --- | --- | --- |
|  |  | Very unlikely | Uncertain | Very likely |
| Protecting oneself (total=15) | To protect oneself |  |  | 10 |
|  | To follow medical advice |  |  | 2 |
|  | Higher protection than single dose |  |  | 2 |
|  | To have both vaccines |  |  | 1 |
| Moving about freely (total=6) | Wanting to travel |  | 1 | 3 |
|  | To gain an immunity passport |  |  | 1 |
|  | Vaccine is a requirement |  |  | 1 |
| Protecting others (total=3) | To protect the wider community |  |  | 3 |
| Lack of trust in authorities and misinformation (total=3) | Lack of trust in government | 1 |  |  |
|  | Lack of trust in science | 1 |  |  |
|  | Conspiracy theory | 1 |  |  |
| Safety concerns (total=3) | Concerns about vaccine side effects |  | 1 |  |
|  | Adverse side effects with first dose | 1 | 1 |  |
| To end pandemic (total=2) | To overcome the pandemic |  |  | 1 |
|  | Vaccine is a civic duty/social responsibility |  |  | 1 |
| Free will (total=2) | Opposing introduction of covid passports |  |  | 1 |
|  | Only want a second dose if it is a free choice |  |  | 1 |

#### Receiving a COVID-19 vaccine

Of 90 participants who had not received any COVID-19 vaccine at T2 (October 2021), 5·6% (95% CI 2·4%, 12·4%, *n*=5) were very likely to have a COVID-19 vaccine, 67·8% (95% CI 57·6%, 77·5%, *n*=61) were very unlikely, and 26·7% (95% CI 18·6%, 36·6%, *n*=24) were uncertain (see Supplementary Figure 2). The modal answer was the minimum value on the intention scale, with 53·3% participants (*n*=48) selecting “0 (extremely unlikely)”.

The most common reasons for not having a COVID-19 vaccine were safety concerns, perceiving the vaccine to be ineffective and preferring natural immunity (Table 5). The main reason behind intention to be vaccinated was to protect oneself.

**Table 5.**
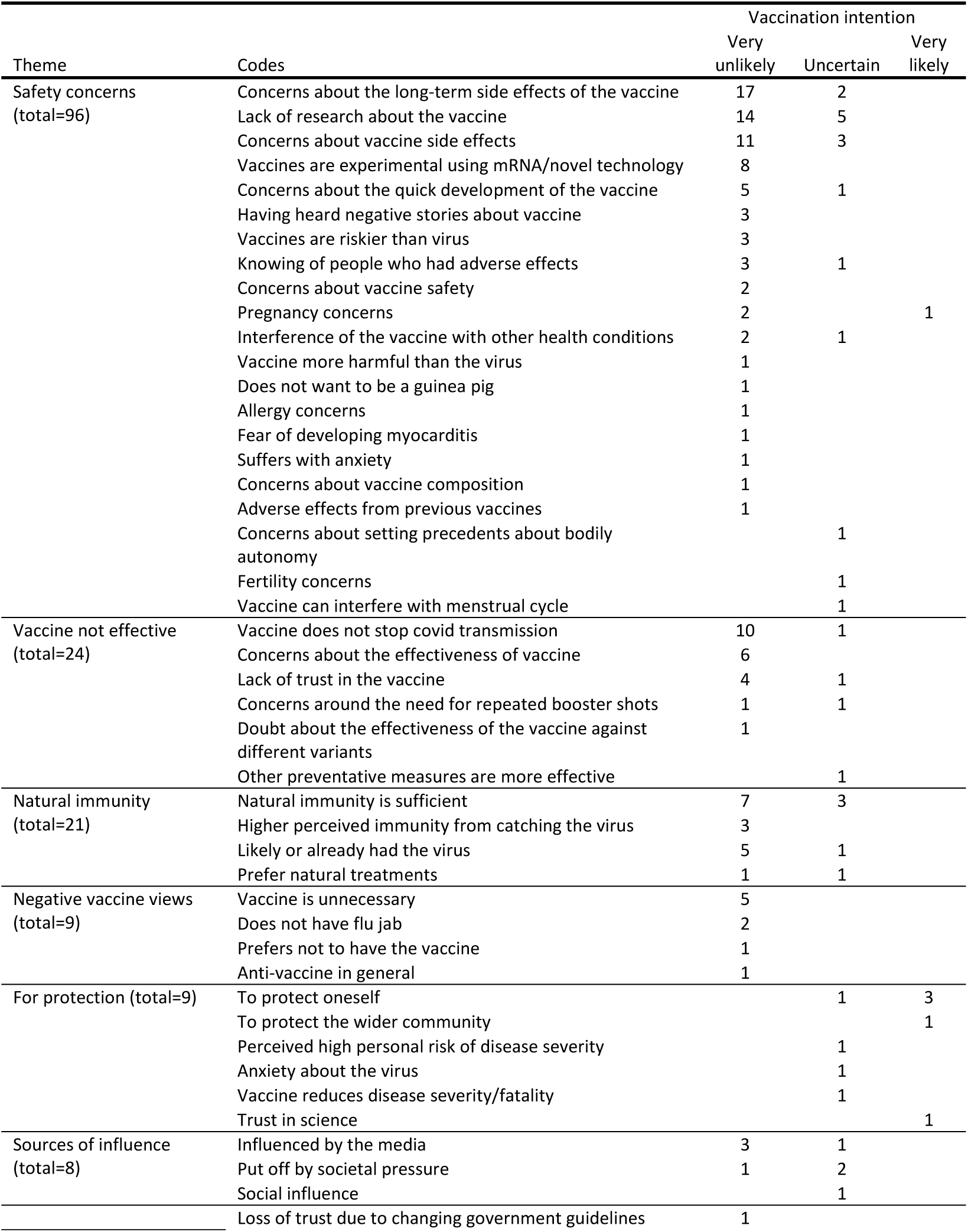
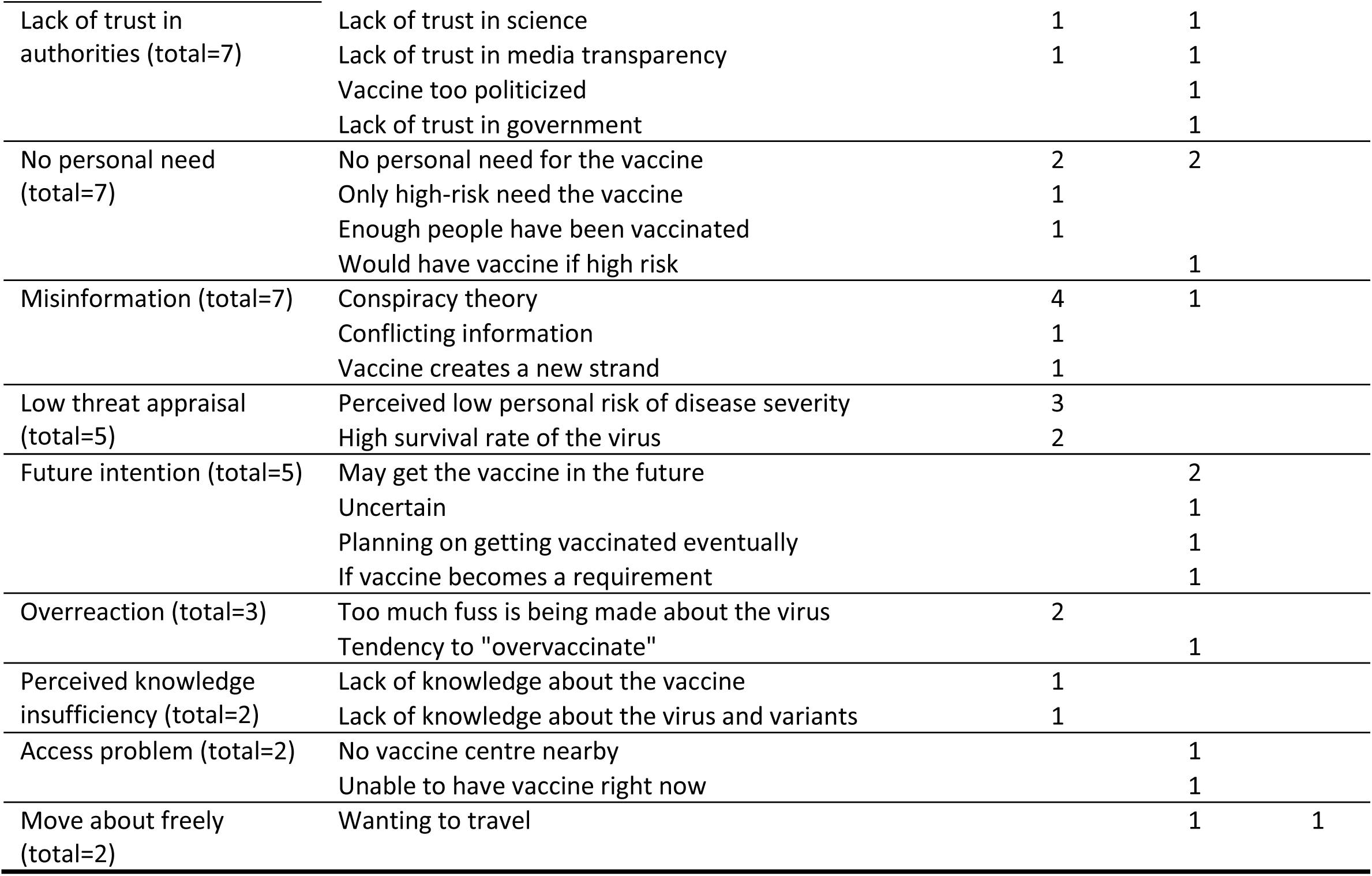
Thematic categorization of codes generated by content analysis of reasons for or against having a COVID-19 vaccine, by vaccination intention. Data show the frequencies with which codes were identified, and themes are presented in descending order of overall frequency.

#### Booster vaccination

Twenty-five participants (2·2%) had already received a COVID-19 booster; these people were excluded from further questions about booster vaccination. Of the remaining 1122 participants, 73·4% (95% CI 70·8%, 75·9%, *n*=823) reported being very likely to receive a COVID-19 booster vaccine if one became available to them; 11·5% (95% CI 9·6% to 13·4%, *n*=129) were very unlikely to do so and 15·2% (95% CI 13·1%, 17·3%, *n*=170) were uncertain. The modal answer was the maximum value on the intention scale, with 59·8% participants (*n*=672) selecting “10 (extremely likely)” (Figure 1).

**Figure 1.**
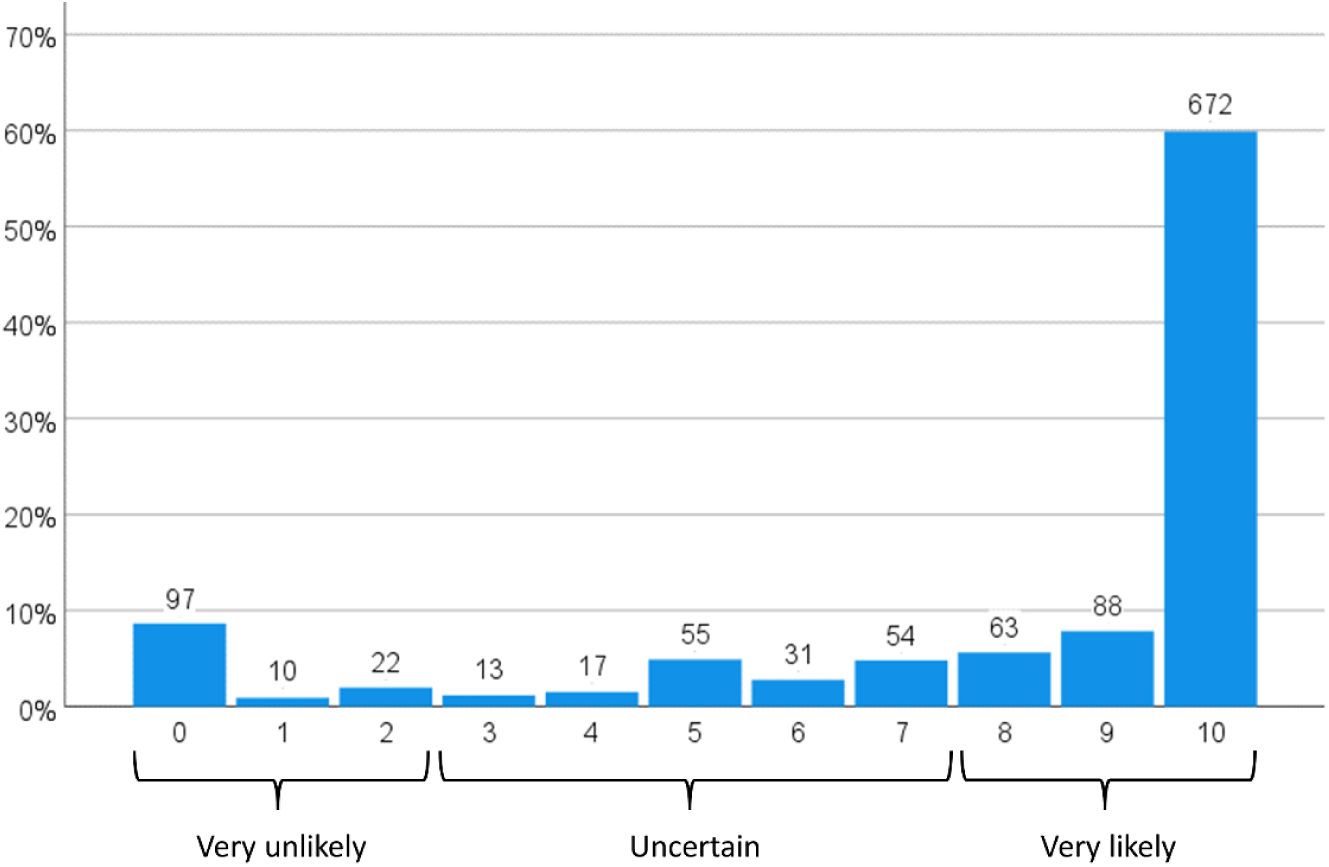
Likelihood of having a COVID-19 booster vaccine, on a scale labelled ‘extremely unlikely’ (0) to 10 ‘extremely likely’ (10), with *a priori* cut-points used to categorize respondents in terms of their booster vaccination intention (*n*=1122).

## Discussion

In our sample, 90% of participants reported having received two COVID-19 vaccines, with a further 2% reporting having had one dose. More people reported having been vaccinated than had previously reported intending to be vaccinated.^1^ This is unusual, as intentions for health behaviours are generally higher than subsequently enacted behaviours.^7^ This is good news for the vaccination campaign in the UK. Our results are not directly comparable to official vaccine statistics, as official figures report on vaccine uptake in those aged 16 years or over (our sample was limited to those aged 18 years and older).^25^ Data stratified by age are available only for England.^26^ Most of our participants reported receiving the AstraZeneca vaccine (57%). Among participants who would have preferred to receive a different vaccine, the overwhelming majority (90%) would rather have received the Pfizer-BioNTech vaccine. This is likely due to the widely publicised associations between adverse effects and the AstraZeneca vaccine.^12^

Vaccine uptake was strongly associated with previous vaccination intention in our study. This is in line with theoretical models of health behaviour (e.g. Protection Motivation Theory^27^), and previous research conducted in other countries.^9-11^ Other psychological factors were also associated with vaccine uptake, namely greater perceived social norms relating to vaccination, greater perceived necessity for vaccination, and greater perceived safety of the vaccine. These factors have also been associated with COVID-19 vaccination intention in other UK studies.^1,2,4^

Of those who had received one dose of the vaccine, intention to have a second dose was high, with the main reason being to protect oneself. In a sample of UK respondents who had not yet decided whether to be vaccinated or who thought they would probably not be vaccinated (conducted October 2020), intention to receive a COVID-19 vaccine was associated with more positive vaccine attitudes and greater perceived safety of vaccination, among other factors.^28^ The pattern in participants who had not received any COVID-19 vaccine was different, with most not intending to be vaccinated in future. The main reasons for this were related to safety concerns surrounding the vaccine, perceptions that the vaccine is not effective, and preferring natural immunity. These factors were also associated with vaccine refusal in previous pandemics.^29^ Taken together, these results suggest that, at a stage where all UK adults had been offered vaccination, those who had started the vaccine programme were likely to complete it, while those who had not received any vaccine were unlikely to do so in the future. Communications should emphasize the safety, effectiveness, and mostly mild side-effects of the COVID-19 vaccine to further increase early uptake.

There was evidence for changes in beliefs and attitudes about COVID-19, COVID-19 vaccination, and general vaccine beliefs between January and October 2021. Participants perceived themselves as being more susceptible to COVID-19, but perceived the illness as less severe. This is likely to reflect the predominant strain circulating in the UK at both timepoints (January 2021: alpha, October 2021: delta). How these changes in beliefs and attitudes affect uptake of booster vaccination remains to be seen. A previous systematic review of vaccine uptake indicated that there was strong evidence that vaccination was associated with perceived susceptibility to infection, but weak evidence for an association between perceived severity of infection; likely because one may consider the likelihood of catching the illness before evaluating its severity.^30^ In October 2021, participants perceived COVID-19 as having a smaller impact on one’s life and there was less emphasis on freedom of restrictions through the vaccine. This may reflect the removal of legal restrictions on mixing in England on 19 July 2021. In contrast to a study of older adults in the UK, which found that worries about unforeseen future effects had increased (between May 2020 and May 2021),^18^ we found that perceptions of COVID-19 vaccine safety increased between January and October 2021. Data collection for the study of older adults was carried out one month after safety concerns about the AstraZeneca vaccine (given to most of the UK population aged 40 years and above) were published in the media.^12^ Contrary to a study conducted in Italy,^17^ we found no evidence for a change in beliefs about commercial profiteering.

Most participants in our study (73.4%) intended to receive a booster vaccine when one became available to them. This is lower than in another UK study (conducted November to December 2021), which found that 8% of participants were unwilling to receive or uncertain about receiving a COVID-19 booster.^31^ This difference may be explained by the fact that their sample comprised only fully vaccinated people. Factors associated with not intending to receive a COVID-19 booster included low levels of stress about catching or becoming seriously ill with COVID-19.^31^

One strength of this study is its longitudinal nature, with participants completing one survey at the start of the COVID-19 vaccine rollout in the UK and another when two doses of the vaccine had been offered to all UK adults. People who completed our T2 survey had higher vaccination intentions than those who did not. Few people indicated that they had received no or just one vaccine dose at T2. Data are self-reported and so are potentially subject to social desirability bias. However, the anonymous nature of the survey should mitigate this.

Official figures show that uptake of the COVID-19 vaccine has been high; this is reflected in self-reported uptake in our sample. Vaccine uptake was associated with higher vaccination intention, perceived safety of vaccination, perceived necessity of vaccination, and social norms for vaccination. Where participants had initiated the vaccine programme, they indicated being likely to complete the vaccination schedule. Where participants had not received any COVID-19 vaccination, they reported being unlikely to begin it. Communications highlighting that severe adverse effects from vaccination are rare and that vaccines are effective may help increase uptake in this group.

## Data Availability

Data are available online at https://osf.io/tehg8/.

## Acknowledgements

The authors are very grateful to the NIHR ARC Covid-19 Research Panel for the Public (now the ARC South London Public Research Panel) for allowing us to present our ideas to them at two separate meetings and for generous and detailed feedback on both occasions. The Panel is chaired by Dr Josephine Ocloo.

## Author contributions

LS, JS, RA, NS, GJR and SMS conceptualized and acquired funding for the study. SMS programmed the survey, curated the data and was responsible for the administration of the project. LS, JS, MC and HD undertook formal analyses. LS wrote the original draft of the manuscript, with support from JS for details of statistical analyses and results. MC, HD, RA, NS, GJR and SMS reviewed and edited drafts.

## Data sharing

Data are available online.^20^

## Supplementary materials

**Supplementary Table 1.**
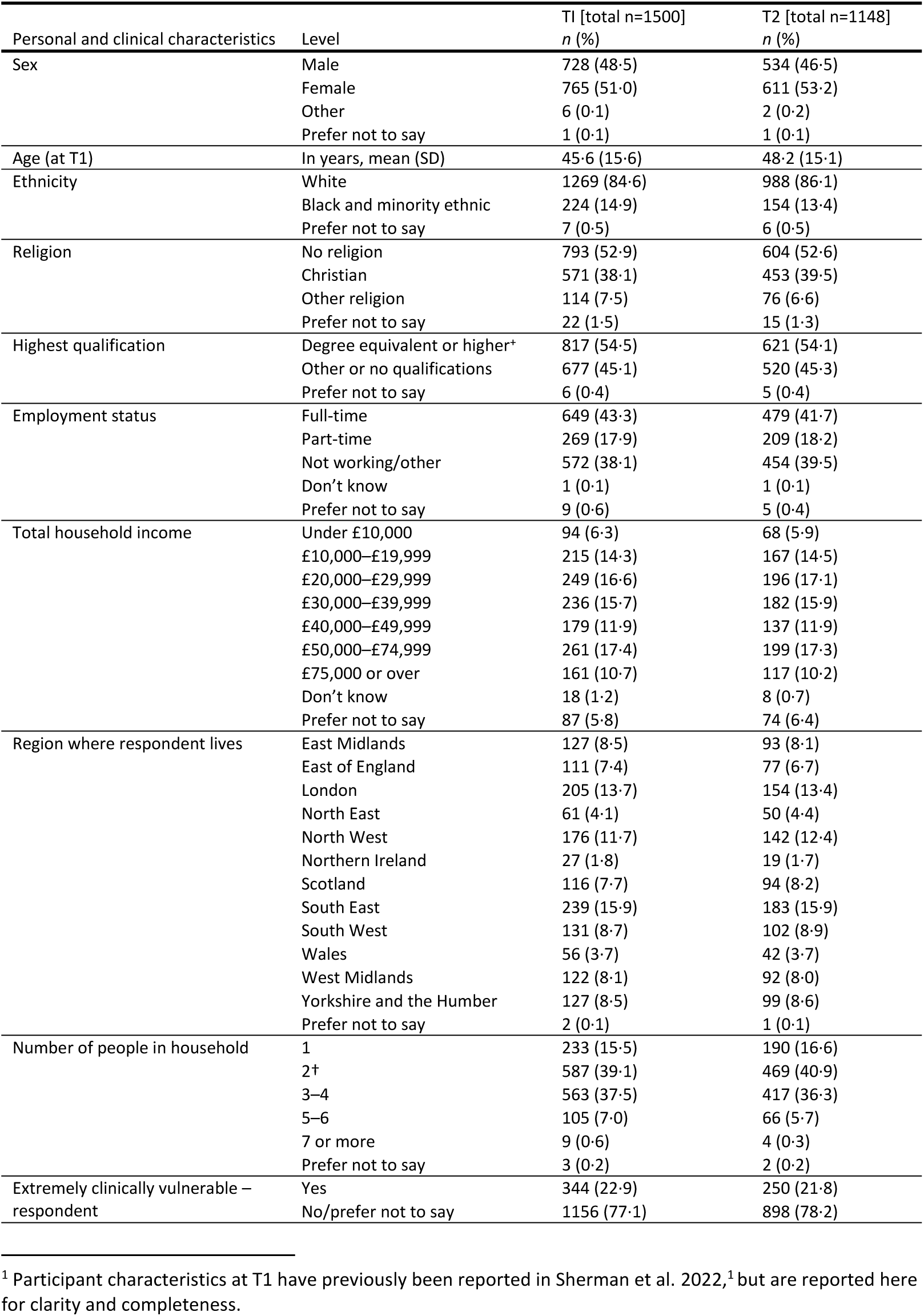
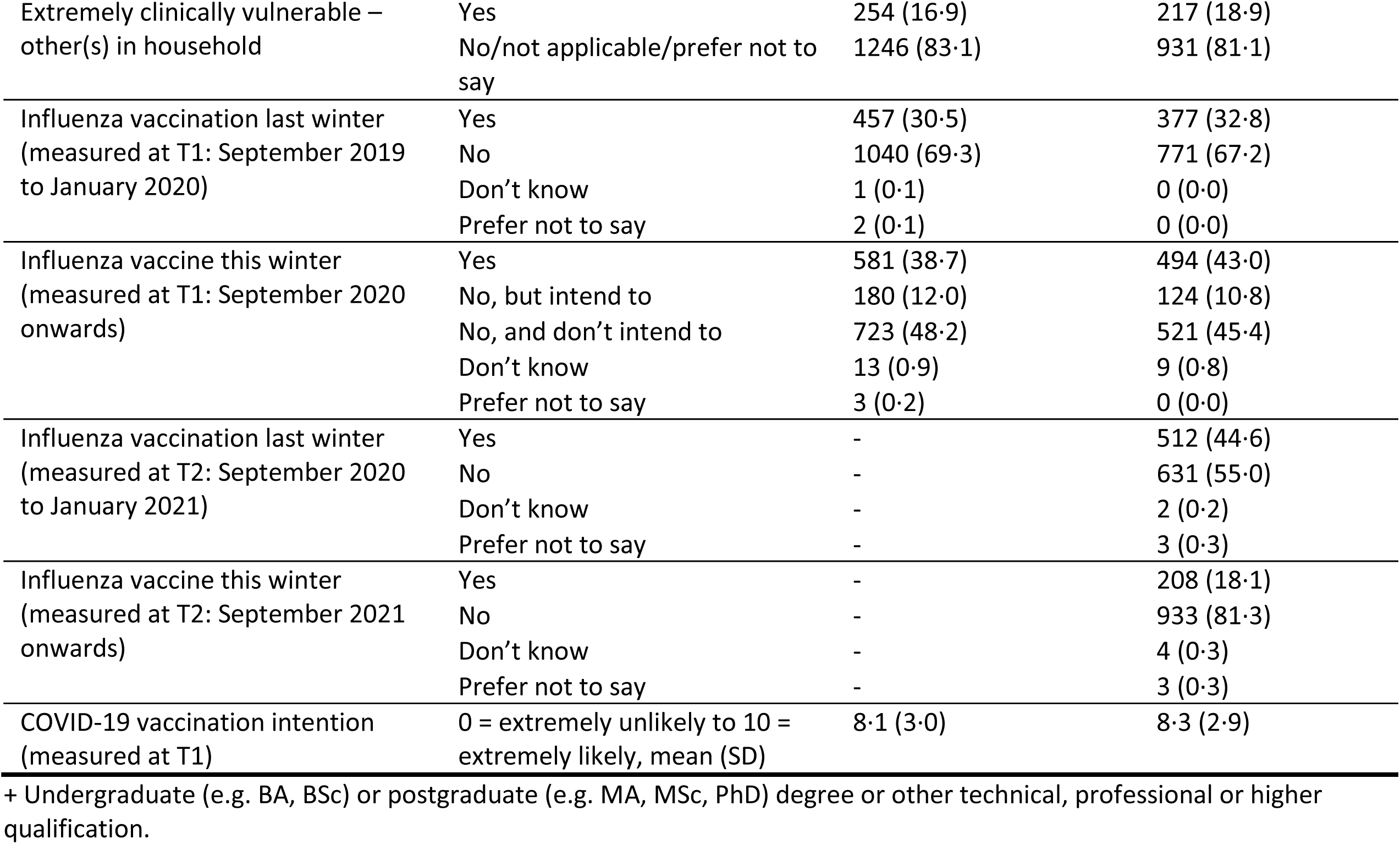
Participant characteristics.^1^

**Supplementary Figure 1.**
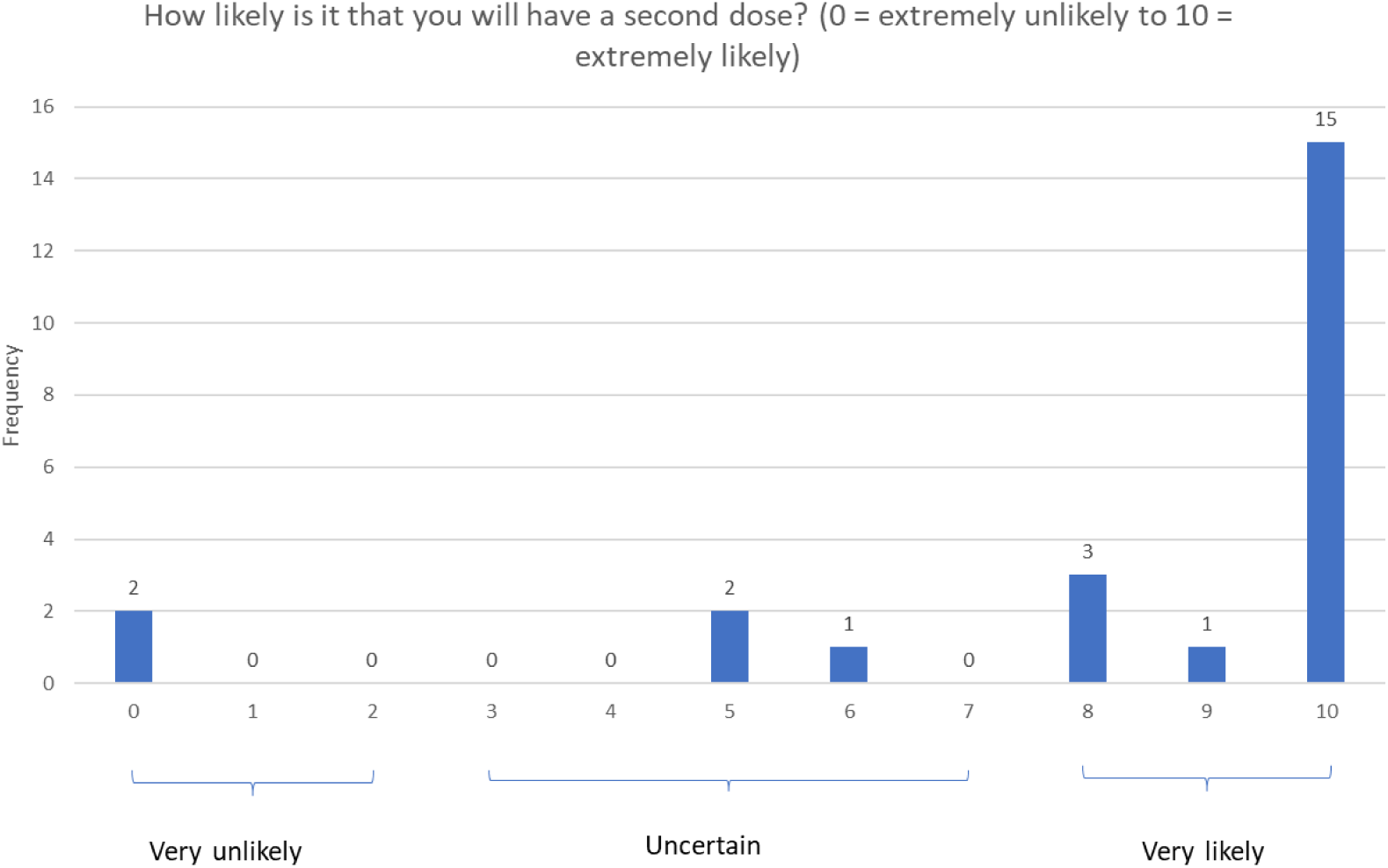
Perceived likelihood of having a second vaccine dose, with *a priori* cut-points used to categorize respondents in terms of their vaccination intention.

**Supplementary Figure 2.**
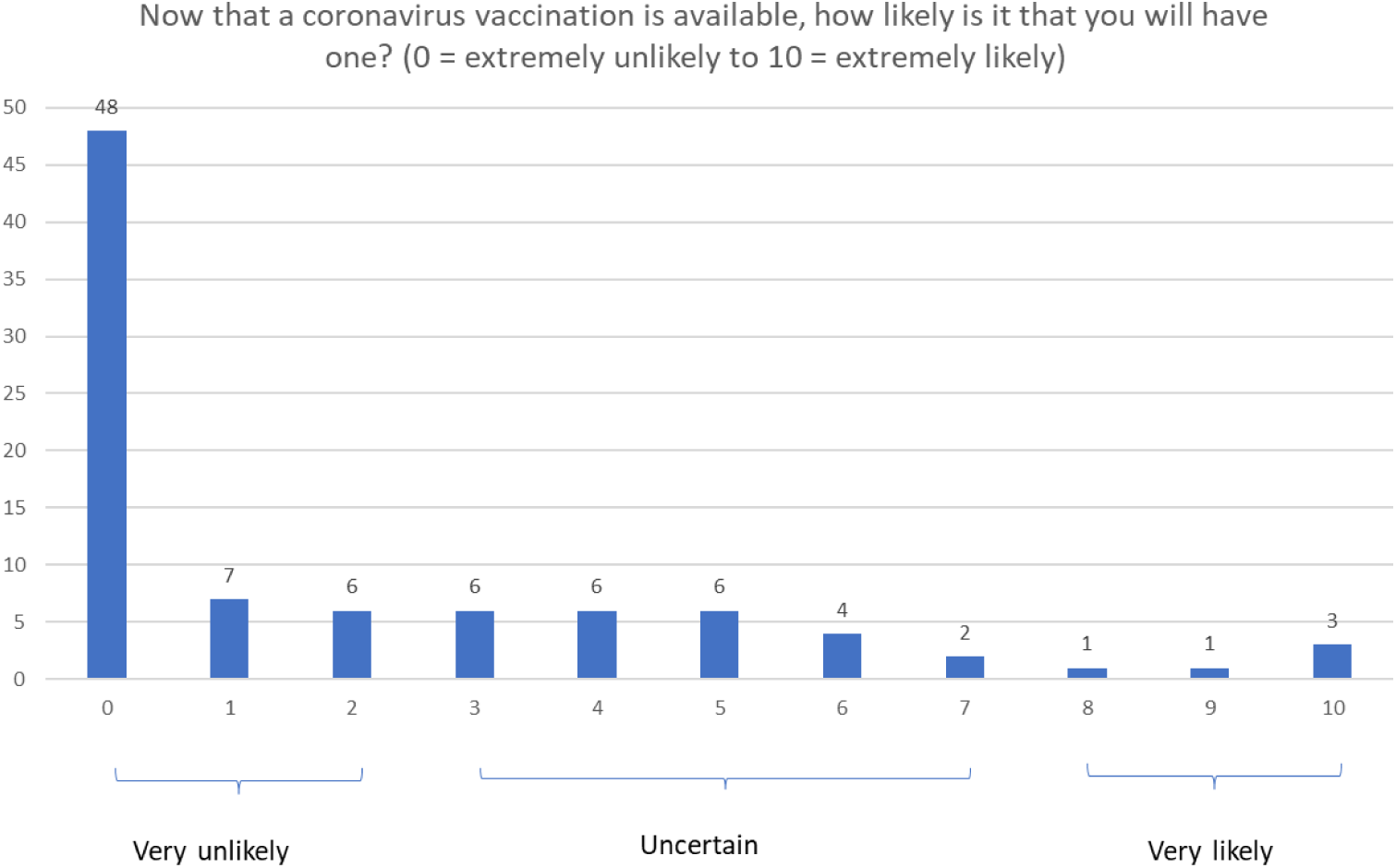
Perceived likelihood of having a COVID-19 vaccine, with *a priori* cut-points used to categorize respondents in terms of their vaccination intention (*n*=90).

## Footnotes

1 Participant characteristics at T1 have previously been reported in Sherman et al. 2022,1 but are reported here for clarity and completeness.

